# Change in Measles Vaccine Coverage and Estimated Impact on Measles Mortality in Low and Middle Income Countries, 2020: An Investigation into Secondary Public Health Effects of the COVID-19 Pandemic Using the Lives Saved Tool

**DOI:** 10.1101/2021.09.07.21263236

**Authors:** Lucy Gregory

**Author notes:** Correspondence concerning this article should be addressed to Lucy Gregory, 1418 Whedbee St, Fort Collins, CO.

## Abstract

With the global COVID-19 pandemic, many public health services were severely disrupted. Estimating the overall health effects of this is difficult as most disease surveillance systems have also been substantially affected during the pandemic. For some diseases, this effect is mitigated by the methods enacted to fight the pandemic, such as use of facial coverings, social distancing and quarantine, but measles is infectious to the degree that this mitigation is likely to be limited. Thus, outbreaks and an increase in global measles mortality are expected. However, the severity of this impact is not yet known.

In early 2020, a study by Roberton and colleagues predicted an additional 12,360 to 37,920 deaths in children under-five worldwide from measles over the coming year based on three potential levels of vaccine coverage reductions ranging from 18.5 to 51.9%. Our study investigates the magnitude of the increase in measles mortality due to decreased vaccine coverage because of COVID-19, based on official estimates of 2020 measles vaccine coverage from WHO/UNICEF released in July 2021. Using the Lives Saved Tool (LiST), an interventions modeling program, we estimated measles mortality for low/middle income countries (LMICs) based on the 2020 WHO/UNICEF estimates of national immunization coverage (WUENIC). Because these calculations use actual reported vaccine coverage, they provide a more accurate picture of measles mortality related to COVID-19 disruptions in 2020.

Using the WUENIC data, LiST predicted fewer additional deaths in 2020 due to decreases in measles vaccine coverage than estimations made by LiST based on Roberton, 2020 due to remarkable recovery efforts by national immunization programmes in the second half of 2020.

---

In 2020, the world saw an upsurgence in measles cases worldwide, but over twenty countries have suspended measles vaccination campaigns in order to cope with the COVID-19 pandemic. It is estimated that over 140 million children did not receive the measles vaccine due to these suspensions. (WHO, 2021) Additionally over 70% of countries reported a disruption to their routine immunization services in 2020 due to the pandemic (WHO Team, 2020). What remains unknown is how this decrease in vaccine coverage translated to measles mortality, especially in low and middle income countries (LMICs).

The Lives Saved Tool, or LiST, is a mathematical modeling program maintained by Johns Hopkins University that estimates the effectiveness in lives saved of community-based public health interventions with a focus on maternal, newborn, and child health. The program is used by researchers, non-profits, advocacy organizations, and governments for public health evaluation, strategic planning, and advocacy. (Stegmuller, 2017) The program was first developed in 2003 as part of the Child Survival Series published in the Lancet and later became open source software through contributions by the Bill and Melinda Gates Foundation. The program contains seventy interventions which have been shown to decrease maternal and child mortality, including measles vaccine coverage. (Walker, 2013) Using the Lives Saved Tool, estimations and predictions can be made for how changes in real or theoretical public health intervention will affect or have affected changes in mortality.

In the early months of 2020, Roberton and colleagues used LiST to estimate the increase in child and maternal mortality due to the indirect effects of the COVID-19 pandemic. Because it was unknown the extent to which various aspects of public health would be affected by the pandemic, the researchers estimated for three conditions of various severity of disruption: decreases in vaccine coverage of 18.5%, 26.9%, and 51.9%. Then, the estimations taken from LiST were compared to baseline coverage to calculate the estimated lives lost due to the indirect effects of the COVID-19 pandemic on 41 public health interventions. For measles specifically, Roberton found that the amount of estimated lives lost worldwide were 1030, 1540, and 3160 per month or 12360, 18480, and 37920 annually for each scenario. It should be remembered that these estimates were made using predicted decreases in vaccine coverage at the beginning of 2020 and not the actual decrease in coverage observed in countries over the year.

The WUENIC (WHO and UNICEF Estimates of National Immunization Coverage) program is an annual collaboration between the WHO and UNICEF. WUENIC uses the best available data and logic-based programming to estimate vaccine coverage in a given year, based on annual coverage totals provided by countries and adjusted to include recent surveys. (Kowalski, 2012) These are generally accepted to be the most accurate estimates of vaccine coverage available and are released yearly in July of the following year. In 2020, WUENIC estimates indicated that an additional 3.7 million children did not receive basic vaccinations in 2020, specifically because of COVID-19 related disruptions. (WHO., & UNICEF, 2021b) As the Roberton estimates were made before the WUENIC data for 2020 were available, we expect that by utilizing the WUENIC estimates for vaccine coverage in conjunction with the Lives Saved Tool, we can create more accurate estimations for the lives lost due to decreased measles vaccine coverage.

The aim of this study is to estimate the number of lives lost in 2020 due to the decrease in measles vaccine coverage using the Lives Saved Tool and data on 2020 vaccine coverage from WUENIC.

## Methods

In the 2020 Roberton study, estimates were made of the number of lives that might be lost due to indirect effects of the COVID-19 pandemic on maternal and child mortality. These estimations were made for three separate scenarios based on potential levels of severity of the secondary effects. Detailed methods are available in the paper but in general coverage reduction was estimated based on the product of reductions in the four domains of workforce, supplies, demand for and access to care.

For vaccinations, the overall reductions in coverage were estimated at 18.5%, 26.9%, 51.9% for the three scenarios. (Roberton, 2020)

Of the countries included in the LiST tool, 118 are considered LMICs by the Organization for Economic Co-operation and Development (OECD). The Gaza Strip was excluded because of incomplete data reporting. Because Roberton did not report on individual countries, we used the LiST to generate estimates of lives lost to measles in 2020 in each of these 117 countries for four scenarios: baseline coverage in 2019, baseline coverage reduced by 18.5%, baseline coverage reduced by 26.9%, and baseline coverage reduced by 51.9%. All of the countries for each of the scenarios were combined to get the total estimated lives lost for that coverage scenario.

### WUENIC

When the 2020 WUENIC Data, (World Health Organization, 2021) were released, an additional 13 countries had no data available for that year. The remaining 104 countries were rerun through the LiST program for mortality with the estimated measles coverage reductions from the Roberton scenarios replaced with the coverage given in the WUENIC data.

**Figure 1:**
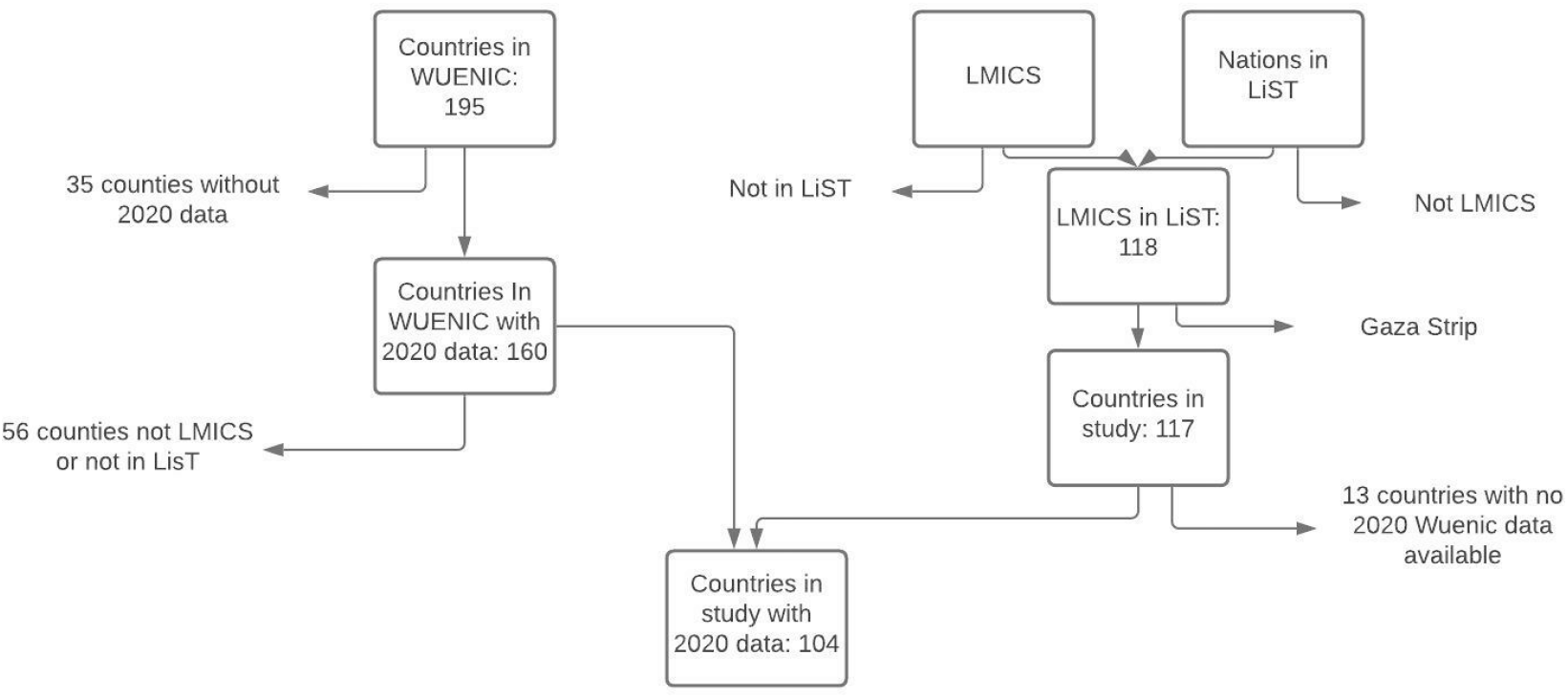
Data flow of Countries Included in Study

### Estimating Additional Deaths

In order to calculate the additional deaths from each of the Roberton scenarios and the reduction of measles vaccination coverage as given by the WUENIC data, the baseline estimate from 2019 was subtracted from the total number of deaths under the four conditions of reduction.

## Results

The 2020 WUENIC data estimates that global measles first-dose vaccine coverage (MCV1) decreased to 84%, the lowest level since 2009. The average change in the 104 countries included in this study was a decrease in coverage of 3.8% from the previous year. WHO/UNICEF estimated that the change from 2019 to 2020 in national MCV1 coverage ranged from a 16% increase (Mexico) to a 30% decrease (Samoa). Between 2019 and 2020, first-dose coverage decreased in 69/104 countries (66.3%), with 35 (33.7%) having decreases greater than 5% and 14 (13.5%) having decreases greater than 10%.

When the WUENIC data was run through the LiST tool, 16 (15.4%) countries had increased mortality in 2020 due to declines in measles vaccine coverage compared to 2019. (Table 1) The number of estimated additional deaths for the year 2020 was 3,555 worldwide, notably lower than the Roberton scenarios which estimated between 16,971 and 45,922 additional deaths.

**Table 1.** Sixteen Countries with Increased Mortality due to Measles from 2019 to 2020 based on WUENIC estimates of MCV1 coverage

| Country | WUENIC 2019 Measles Coverage | WUENIC 2020 Measles Coverage | 2019 Estimated Measles Mortality (Baseline) | Robertson 1 (Low) Mortality | Robertson 2 (Med) Mortality | Robertson 3 (High) Mortality | Estimated 2020 Measles Mortality from WUENIC data |
| --- | --- | --- | --- | --- | --- | --- | --- |
| Angola | 51 | 44 | 400 | 425 | 436 | 469 | 418 |
| Benin | 68 | 65 | 155 | 171 | 178 | 200 | 159 |
| Congo | 73 | 68 | 367 | 419 | 442 | 512 | 386 |
| Côte d'Ivoire | 76 | 70 | 2007 | 2353 | 2508 | 2974 | 2154 |
| Gabon | 62 | 53 | 139 | 149 | 153 | 167 | 147 |
| India | 95 | 89 | 6812 | 13678 | 15326 | 20209 | 9540 |
| Indonesia | 88 | 76 | 3063 | 4043 | 4487 | 5809 | 3784 |
| Liberia | 68 | 61 | 1624 | 1791 | 1867 | 2093 | 1717 |
| Madagascar | 64 | 59 | 153 | 168 | 175 | 196 | 160 |
| Mali | 70 | 62 | 232 | 256 | 267 | 301 | 247 |
| Mauritania | 78 | 72 | 98 | 112 | 118 | 136 | 104 |
| Philippines | 73 | 72 | 310 | 359 | 381 | 447 | 314 |
| Samoa | 87 | 57 | 2 | 2 | 3 | 3 | 3 |
| Sudan | 90 | 86 | 843 | 1024 | 1107 | 1353 | 887 |
| Togo | 75 | 69 | 79 | 89 | 94 | 107 | 84 |
| Ukraine | 93 | 85 | 17 | 24 | 28 | 37 | 21 |

Three nations (Djibouti, Montenegro, Suriname) had coverage reductions equal to or greater than the lowest scenario (18.5%) in Roberton; two nations, Montenegro and Suriname, had coverage reductions equal to or greater than the middle scenario (26.9%). No nations had coverage decreases equal or exceeding the third scenario (51.9%). Samoa was the only nation where the WUENIC data indicated a number of additional deaths that reached any of the Roberton scenarios. On the other end of the spectrum, Bangladesh, DRC, Eritrea, Nigeria, Pakistan, and Somalia all had many fewer estimated deaths using WUENIC data than was calculated in the Roberton scenarios. In the Roberton predictions, measles mortality in these nations was highly sensitive to changes in coverage, but the estimated coverage reductions for these countries from the WUENIC data were much less than predicted.

## Discussion

While much of the world is focused on the direct mortality from the Covid 19 pandemic, secondary mortality from disrupted essential healthcare services and diverted healthcare resources is harder to quantify and therefore overlooked and deprioritized. Vaccination, and particularly measles vaccine, is one of the most effective and cost-effective childhood survival interventions. Improved measles vaccination coverage has saved an estimated over 25 million lives since 2000 (Patel, 2020), and any erosion of vaccine coverage can rapidly lead to outbreaks and large numbers of childhood deaths. Using 2020 WUENIC data, which reflected the tremendous efforts of vaccination programs to sustain measles coverage, we found that estimates of excess measles mortality were lower than predicted early on in the pandemic. While the numbers generated by the LiST modelling tool are only estimates, they provide a framework by which the impact of disruptions and interventions can be evaluated by policy makers and the public.

Where previous studies had to rely on a range of possible scenarios, we were able to generate a more accurate picture of the COVID 19 pandemic’s impact on measles mortality by using the actual data provided by the WUENIC report. Compared to the estimated deaths generated by the Roberton study, overall deaths using the WUENIC data were 74.3% lower than even the Roberton best-case scenario. This suggests that, while still existent, increased measles mortality due to secondary effects of the pandemic was lower than expected. Data released by UNICEF and WHO (Unicef, & WHO, 2021) indicate that while vaccine coverage did decrease by up to 60% in some regions in the first 6 months of 2020, there was a major recovery effort in the latter half of the year across all regions that appears to have been effective in mitigating the initial drop. Countries in Southeast Asia and the Eastern Mediterranean were most affected by the vaccination decreases, as were the lower income countries which suffered a disproportionate drop in coverage compared to higher income countries. It is important to remember, however, that even a relatively small drop in coverage corresponds to over 3 million more children unvaccinated against measles (UNICEF, & WHO, 2021).

The Roberton modelling did not account for national variations in coverage; their scenarios assumed an equal percentage decrease in coverage for all included countries. In reality, the extent to which the pandemic disrupted normal public health activity varied widely between nations. In Pakistan, measles vaccine coverage per WUENIC estimates actually increased by two points in 2020, leading to an estimated 300 fewer deaths than even pre-COVID-19 modeling would have predicted. One reason why the estimated additional measles deaths are lower than predicted is that vaccine-coverage reduction in Sub-Saharan Africa was minimal in 2020. Many nations in that region are historically major contributors to global measles mortality, whereas many of the nations that contributed most to the estimated additional deaths in this study were in areas with high coverage disruption, such as South and Southeast Asia, which traditionally have a smaller percentage of total measles deaths (Patel, 2020).

Public health estimations done by modeling tools like LiST, from national or local disease reporting systems, or by committees of experts, are always, inherently, inexact. Exact numbers on mortality rates, coverage, lives saved by an intervention, etc, are impossible to generate due to uncertainty in the underlying data. In addition, the conditions of 2020 and the predictive nature of LiST added further uncertainty around estimates of the exact mortality due to decreased measles coverage.

The WUENIC data has inherent inaccuracies. WUENIC estimates are created by compiling coverage reported by each nation, and then adjusting that data in order to incorporate other data sources like recent coverage surveys and vaccine stockouts (Pan American Health Organization, 2020). However, in the year 2020, due to disruptions caused by the COVID-19 pandemic, only 160 countries submitted reports, compared to 195 in 2019. (WHO, & UNICEF, 2021a). In addition, the accuracy of the data in many countries depends heavily on periodic coverage surveys to confirm or adjust administrative coverage. The further out current estimates are from survey data the more uncertainty exists, and WUENIC estimates are frequently carried forward from one year to another in the absence of additional recent data, a situation exacerbated by the pandemic when coverage surveys were also disrupted. In addition, survey data relies to a variable degree on caregiver recall of vaccine receipt which can be another source of uncertainty and bias for the estimates. Some subjective decisions need to be also made in the process of generating WUENIC estimates (Kowalski, 2012)

At the same time that the 2020 WUENIC estimates were released, another group led by the Institute for Health Metrics and Evaluation (IHME) released modeled estimates of measles vaccine coverage based on administrative and mobility data and a measure of vaccine coverage disruption (Causey, 2021). This group estimated that MCV1 coverage had declined by 7.9% from 2019, much higher than the estimated 2% decrease apparent in the WUENIC estimates. This is equivalent to almost 3 times as many children missing out on measles vaccination due to the COVID-19 pandemic than the WUENIC estimate (8.9 million versus 3 million). This projection used data for vaccine coverage disruption that primarily came from the first half of 2020 and may not therefore fully reflect the recovery of vaccination in the second half of the year, therefore overestimating the total annual decrease in MCV1 coverage. However if the true coverage were closer to these estimates, the numbers of estimated measles deaths would likely be significantly higher than those in our study. Because the paper by Causey and colleagues only provides estimated changes in coverage by region rather than country, it is not impossible to use the LiST tool to directly compare expected deaths from these estimates to those from WUENIC data.

Measles is an outbreak-prone disease, making year-to-year case levels and predicted deaths per country difficult to predict. However, the data for under-five mortality used by LiST only goes until the year 2017, and then is extrapolated from there. This means that the per cause mortality, one of the major factors the tool uses to estimate lives lost to a particular cause, for each country after 2017 may be inaccurate.

## Conclusion

While many nations saw increased mortality due to measles in 2020, this increase appears to be less than predicted at the beginning of the pandemic, due in part to the strong recovery efforts of public health workers in LMICs in the latter half of 2020. However, coverage and change in coverage varied vastly between countries during 2020 and preliminary data suggests this trend is likely to continue in 2021. Long-term improvements in measles immunization likely provide some level of immunity against short lapses in coverage, but will not stand up in the face of prolonged disruptions, particularly given ongoing disruption to supplementary immunization activities traditionally used in many countries to fill immunity gaps among children missed by routine immunization services. Low mortality numbers should not falsely reassure policy makers into not prioritizing immunization recovery efforts as they are likely to be transitory in the absence of robust catch-up activities.

## Data Availability

Raw data were generated from WUENIC results. Derived data supporting the findings of this study are available from the corresponding author LG on request.

https://apps.who.int/immunization_monitoring/globalsummary/timeseries/tswucoveragedtp3.html

## Acknowledgements

We thank Laura L. Hammitt, MD, Associate Professor, Johns Hopkins Bloomberg School for Public Health for her thoughtful assistance with editing this paper, and Jerry Thomas, PhD for his encouragement to begin and continue this project.

